# Artificial intelligence and omics-based autoantibody profiling highlights autoimmunity targeting ligand-receptor interaction in dementia

**DOI:** 10.1101/2024.09.20.24313547

**Authors:** Kazuki M Matsuda, Yumi Umeda-Kameyama, Kazuhiro Iwadoh, Masashi Miyawaki, Mitsutaka Yakabe, Masaki Ishii, Sumito Ogawa, Masahiro Akishita, Shinichi Sato, Ayumi Yoshizaki

**Affiliations:** Department of Dermatology, The University of Tokyo Graduate School of Medicine, Tokyo, Japan; Department of Geriatric Medicine, The University of Tokyo Graduate School of Medicine, Tokyo, Japan; Tokyo Metropolitan Institute for Geriatrics and Gerontology, Tokyo, Japan; Department of Clinical Cannabinoid Research, The University of Tokyo Graduate School of Medicine, Tokyo, Japan

**Author notes:** **Corresponding author** Ayumi Yoshizaki, MD, PhD Department of Dermatology and Department of Clinical Cannabinoid Research, The University of Tokyo Graduate School of Medicine, 7-3-1, Hongo, Bunkyo-ku, Tokyo, Japan, 1138655.

## Abstract

Dementia is a neurodegenerative syndrome marked by the accumulation of disease-specific proteins and immune dysregulation, including autoimmune mechanisms involving autoantibodies. Current diagnostic methods are often invasive, time-consuming, or costly. This study explores the use of proteome-wide autoantibody screening (PWAS) for noninvasive dementia diagnosis by analyzing serum samples from Alzheimer’s disease (AD), dementia with Lewy bodies (DLB), and age-matched cognitively normal individuals (CNIs). Serum samples from 35 subjects were analyzed utilizing our original wet protein arrays that covers approximately 90% of human transcriptome, revealing elevated gross autoantibody levels in AD and DLB patients compared to CNIs. A total of 229 autoantibodies were differentially elevated in AD and/or DLB, effectively distinguishing between patient groups. Machine learning models showed high accuracy in classifying AD, DLB, and CNIs. Gene ontology analysis highlighted autoantibodies targeting neuroactive ligands/receptors in AD and lipid metabolism proteins in DLB. Notably, autoantibodies targeting neuropeptide B (NPB) and adhesion G protein-coupled receptor F5 (ADGRF5) showed significant correlations with clinical traits including Mini Mental State Examination scores, suggesting a role in dementia pathogenesis. The study demonstrates the potential of PWAS and AI integration as a noninvasive diagnostic tool for dementia, uncovering biomarkers that could enhance understanding of disease mechanisms. Limitations include demographic differences, small sample size, and lack of external validation. Future research should involve longitudinal observation in larger, diverse cohorts and functional studies to clarify autoantibodies’ roles in dementia pathogenesis and their diagnostic and therapeutic potential.

## Introduction

Dementia is a complex neurodegenerative syndrome affecting millions worldwide. Early diagnosis is crucial for timely intervention, yet many current diagnostic methods are either invasive, time-consuming, or expensive. For instance, psychological assessments require significant time and concern to patients themselves, cerebrospinal fluid examination is invasive, and amyloid positron emission tomography (PET) is costly. Consequently, there is a pressing need for a simpler, noninvasive, and cost-effective diagnostic method for dementia.^1–3^

Pathologically, dementia is marked by the aggregation of disease-specific proteins in the brain.^4^ While the pathogenic role of abnormal protein deposition in dementia is well-established, the precise mechanisms behind the initiation and progression of neurodegeneration remain unclear. Meanwhile, emerging evidence has highlighted the role of immune dysregulation in dementia’s pathogenesis. Genome-wide association studies have identified common genetic variations in immune system processes that are associated with neurodegenerative diseases such as Alzheimer’s disease (AD), frontotemporal dementia (FTD), and Parkinson’s disease dementia.^5–7^

Autoimmune mechanisms are gaining recognition as a key factor in the pathophysiology of dementia.^8–10^ Autoantibodies—self-reactive antibodies produced by B cells—play a role in immune tolerance and homeostasis.^11^ However, due to various genetic and environmental factors, the ability to distinguish “self” from “non-self” deteriorates, leading autoantibodies to trigger and sustain inflammatory processes that cause tissue damage.^12,13^ Autoantibodies have been detected in both blood and cerebrospinal fluid of patients with various forms of dementia, including autoimmune dementia and neurodegenerative dementias such as AD, FTD, vascular dementia (VD), and dementia with Lewy bodies (DLB).^14–18^ Autoimmune dementia is characterized by progressive cognitive decline with an early onset, atypical clinical presentation, rapid progression, the presence of neural antibodies, cerebrospinal fluid inflammation, brain changes in MRI atypical for neurodegenerative diseases, and a good response to immunotherapy.^19^ Various neural autoantibodies have been frequently identified in individuals with progressive cognitive decline, targeting cell surface proteins such as the N-methyl-D-aspartate receptor, gamma-aminobutyric acid B receptor, alpha-amino-3-hydroxy-5-methyl-4-isoxazolepropionic acid receptor, leucine-rich glioma inactivated protein 1, dipeptidyl-peptidase protein-like 6, and transcobalamin receptor.^20–22^ However, there is an overlap in the neural autoantibody profiles between autoimmune dementia and neurodegenerative dementias like FTD and DLB, necessitating further research to clarify disease specificity.^20^

AD, one of the most well-known forms of dementia, is characterized by the accumulation of amyloid plaques and neurofibrillary tangles in the brain.^23^ Autoantibodies targeting amyloid-β (Aβ), tau, neurotransmitters, and microglia have been reported in AD patients.^24,25^ Specifically, autoantibodies against Aβ are decreased in AD patients,^26,27^ suggesting a protective role against Aβ toxicity,^28,29^ in line with clinical efficacy of lecanemab, a humanized monoclonal antibody targeting Aβ soluble protofibrils.^30^ Additionally, increased levels of autoantibodies against glutamate,^31^ oxidized low-density lipoproteins,^32^ glial markers such as GFAP and S100B,^33^ and receptors for advanced glycosylation end products have been observed in AD patients’ serum or cerebrospinal fluid.^34^ DLB is another progressive neurodegenerative disorder characterized by the presence of Lewy bodies—abnormal aggregates of the protein alpha-synuclein—in the brain.^35^ Autoantibodies against alpha-synuclein, Aβ, myelin oligodendrocyte glycoprotein, myelin basic protein, and S100B, have been identified in some DLB patients.^36,37^ Autoantibodies have been detected even in patients with mild cognitive impairment (MCI), indicating a potential role in disease progression.^38–40^ Despite the discovery of autoantibodies related to various forms of dementia pathology, further research is needed to assess their potential as diagnostic or prognostic biomarkers and their utility in developing effective immunotherapies for dementia.^28^

One promising approach is the use of protein microarrays for autoantibody profiling, which could help identify novel autoantibodies for diagnosing and monitoring MCI and dementia.^39,40^ In this pilot study, we utilized a proteome-wide autoantibody screening (PWAS) technique employing wet protein arrays (WPAs) that cover approximately 90% of the human transcriptome.^41,42^ This method has previously been used to develop multiplex measurements for disease-related autoantibodies,^43,44^ identify clinically relevant novel autoantibodies,^45–48^ and investigate epitope spreading during disease progression.^49^ We have successfully applied this technique to a variety of inflammatory disorders, including systemic sclerosis,^47^ and identified autoantibodies to membranous antigens like G protein-coupled receptors (GPCRs) using machine learning approaches.^48^ In this study, we applied PWAS to serum samples from patients with AD or DLB and age-matched cognitively normal individuals (CNIs) to elucidate the autoantibody landscape in dementia. Our goal was to identify clusters of autoantibodies that may contribute to the pathophysiology of dementia, by integration of artificial intelligence (AI) and omics-based approach. This research aims to uncover novel biomarkers and enhance our understanding of dementia’s pathogenesis.

## Results

### Demographic and clinical characteristics

Serum samples from 35 subjects, including 18 patients with AD, 8 patients with DLB, and 9 CNIs were served for PWAS utilizing WPAs. The baseline demographics across the three groups were similar, except that the proportion of females was highest in the AD group and lowest among CNIs (**Extended Table 1**). The proportion of females in the AD, DLB, and CNI groups were 82.4%, 62.5%, and 33.3%. The Hasegawa’s Dementia Scale-Revised (HDSR) scores for the each group were 19.9±5.6, 22.1±5.6, and 27.9 ± 2.0, respectively, while the Mini Mental State Examination (MMSE) scores were 20.2±3.9, 21.1±6.6, and 28.9±1.4.

### Sum of autoantibody levels

We defined the sum of autoantibody levels (SAL) as the total serum concentration of all autoantibodies measured in our PWAS. Although not statistically significant, SAL was higher in patients with AD and DLB compared to CNIs (**Figure 1A**). This trend persisted across all age groups (**Extended Figure 1A**) and was relatively higher in females than in males (**Extended Figure 1B**).

**Figure 1.**
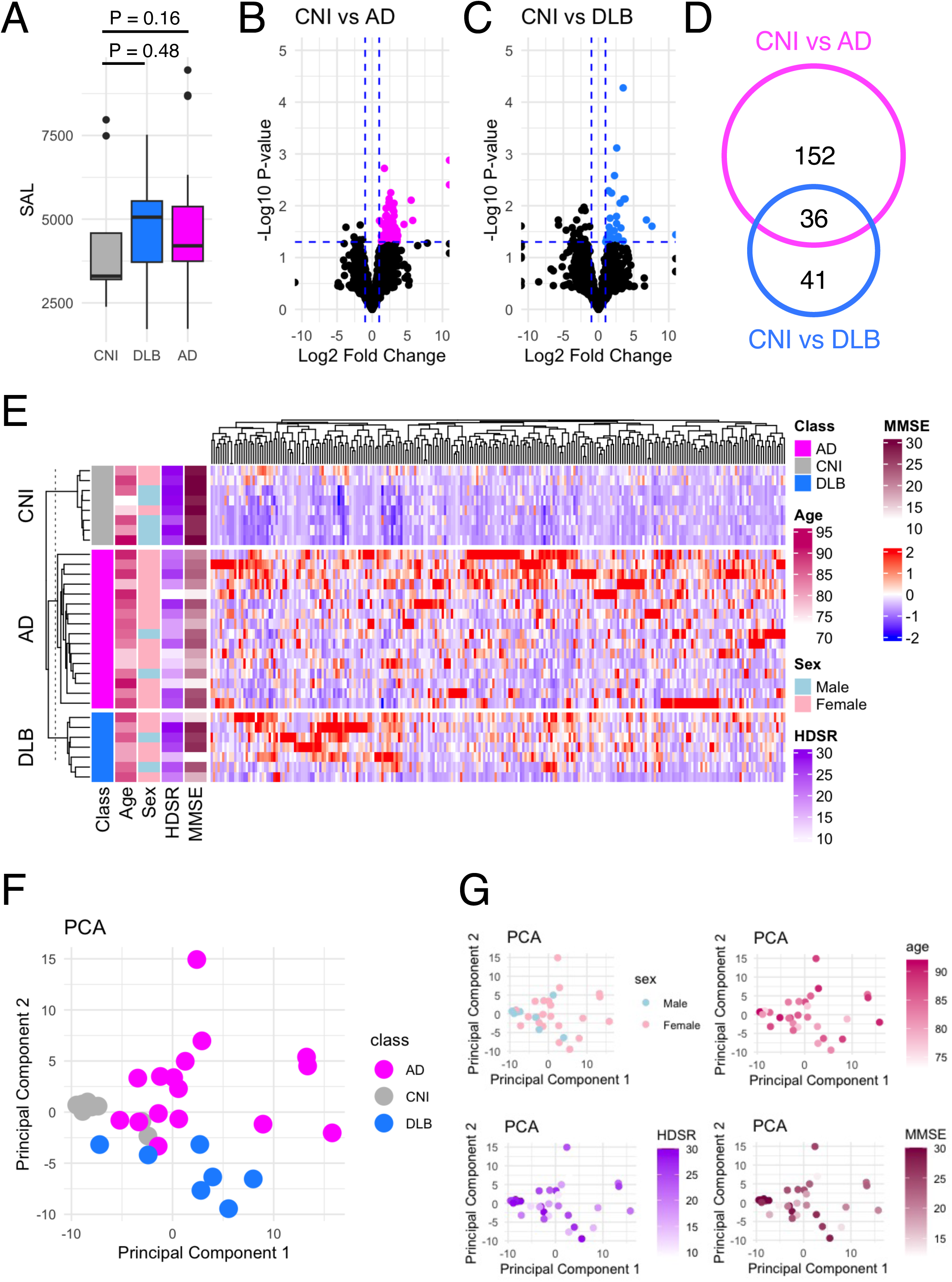
Autoantibodies differentially elevated in dementia. **(A)** The SAL in AD, DLB, and CNI. **(B)** Volcano plot that shows autoantibodies differentially elevated in AD compared to NCI. The vertical dash line indicates P = 0.05. The horizontal dash line indicates fold change = ±2. **(C)** Volcano plot that shows autoantibodies differentially elevated in DLB compared to NCIs. The vertical dash line indicates P = 0.05. The horizontal dash line indicates fold change = ±2. **(D)** Venn diagram that illustrates the inclusion relationship between autoantibodies differentially elevated in AD and/or DLB. The vertical dash line indicates P = 0.05. The horizontal dash line indicates fold change = ±2. **(E)** Heat map that shows the serum levels of 229 autoantibodies differentially elevated in AD and/or DLB. **(F)** PCA of 229 autoantibodies differentially elevated in AD and/or DLB. In the scatter plot, individual subjects as points. **(G)** PCA plots colored by sex, age, HDSR, and MMSE.

### Identification of differentially elevated autoantibodies

Next, we focused on identifying autoantibodies with serum levels significantly elevated in AD (**Figure 1B**) and/or DLB (**Figure 1C**) compared to CNIs. This analysis revealed 188 autoantibodies elevated in AD and 77 in DLB, with 36 overlapping between the two conditions (**Figure 1D**), totaling 229 distinct items (**Figure 1E**). Using these autoantibodies, we performed principal component analysis (PCA), which effectively differentiated AD patients, DLB patients, and CNIs (**Figure 1F**), regardless of sex, age, or cognitive impairment severity as measured by HDSR and MMSE (**Figure 1G**).

### AI-based 2-class classification

To identify which of the 229 autoantibodies were most strongly associated with disease status, we employed 14 different machine learning frameworks. Logistic regression with normalization or standardization, along with support vector machines (SVM) under similar conditions, achieved an area under the receiver operating characteristic curve (ROC-AUC) exceeding 0.96, indicating near-perfect accuracy in distinguishing AD patients from others (**Table 1**). We identified the top 10 features from these four models (**Figure 2A**), assessed their overlap (**Figure 2B**), and analyzed the serum levels of 12 autoantibodies highlighted in more than two frameworks (**Figure 2C**).

**Figure 2.**
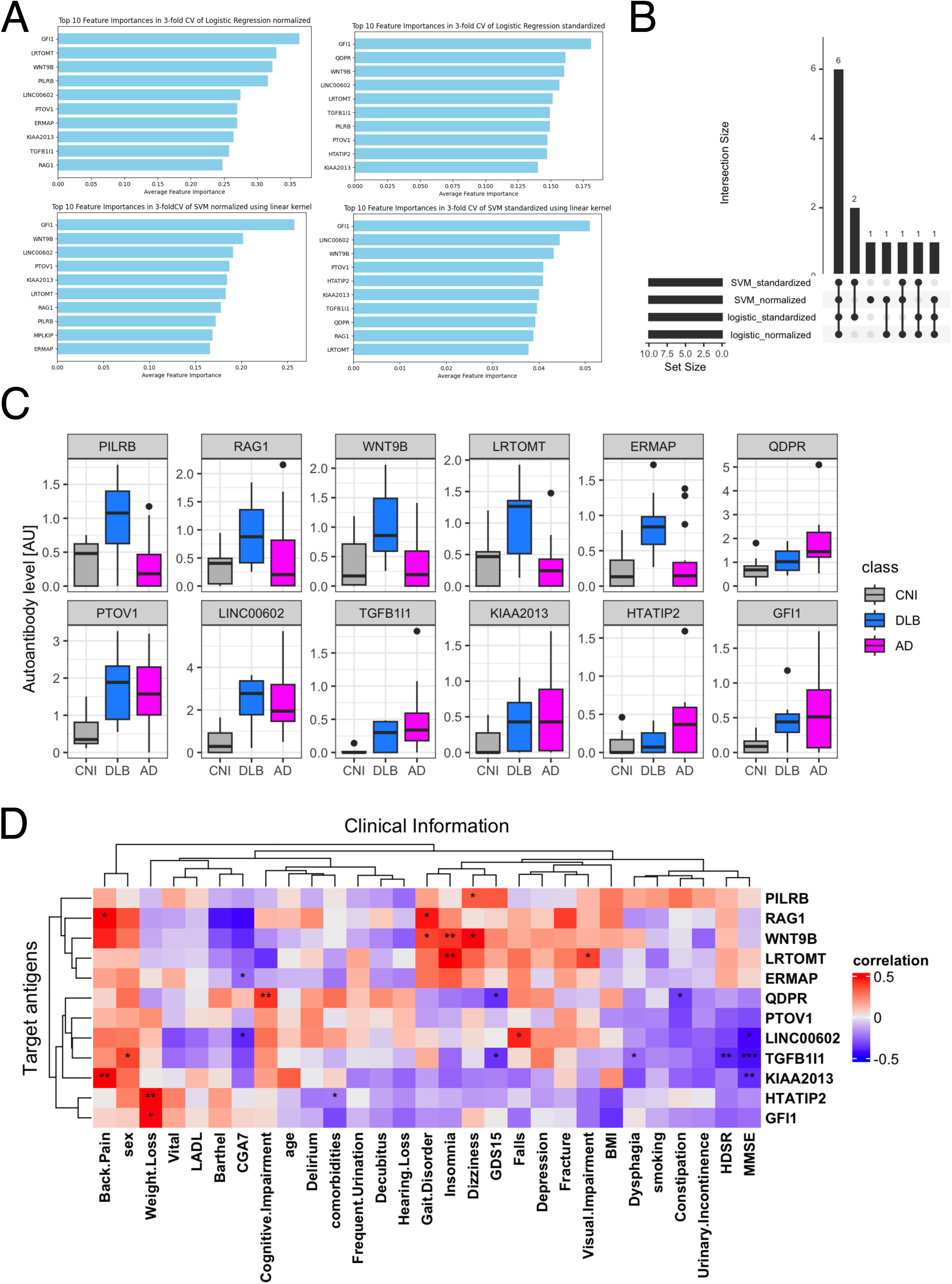
Autoantibodies highlighted in 2-class classification tasks by AI. **(A**) Autoantibodies that were mostly highlighted according to feature importance by Logistic regression and SVM with standardization or normalization. **(B**) UpSet plot shows the inclusion relationship of autoantibodies highlighted by the four machine learning frameworks. **(C)** Box plots describe the serum levels of autoantibodies highlighted by more than two frameworks in AD, DLB, and CNI. **(D)** Heatmap illustrates correlation between autoantibodies highlighted in machine learning analysis and demographic and clinical characteristics of dementia. *: P < 0.05, **: P < 0.01. P values were calculated by Spearman’s correlation test.

Next, we examined the relationship between these 12 autoantibodies and clinical traits (**Figure 2D**). This analysis revealed a significant correlation of serum levels of autoantibodies targeting proteins encoded by *TGFB1I1* and *KIAA2013* with HDSR scores. However, a database search, utilizing the Human Protein Atlas,^50^ indicated that these two genes are not specifically expressed in the central nervous system (data not shown). Although serum levels of anti-TGFB1I1 antibodies were significantly associated with sex, trends in the distribution of these autoantibodies among three groups were generally similar between both sex (**Extended Figure 2A**). To evaluate cross-reactivity, we performed a correlation analysis on these 12 autoantibodies. Those with moderate to high correlations (Spearman’s r > 0.5) underwent sequence alignment and identity analysis. The correlation matrix (**Extended Figure 2B**) revealed five correlated autoantibodies, and sequence analysis showed that all proteins shared less than 25% identity (**Extended Figure 2C**). Additionally, we investigated the prevalence of these highlighted autoantibodies across a broader spectrum of human disorders using the aUToAntiBody Comprehensive Database (UT-ABCD).^47^ Most of these autoantibodies were found to be non-specifically elevated in various pathological conditions (**Extended Figure 2D**).

### AI-based 3-class classification

We also explored multi-class classification among AD, DLB, and CNI by training deep neural networks with five hidden layers using the 249-dimensional autoantibody profiles. The optimal number of epochs was determined based on the accuracy and loss trajectories (**Figure 3A**). This approach resulted in high accuracy, with ROC-AUC values reaching up to 0.95 (**Figure 3B**), as well as high precision and recall (**Figure 3C**).

**Figure 3.**
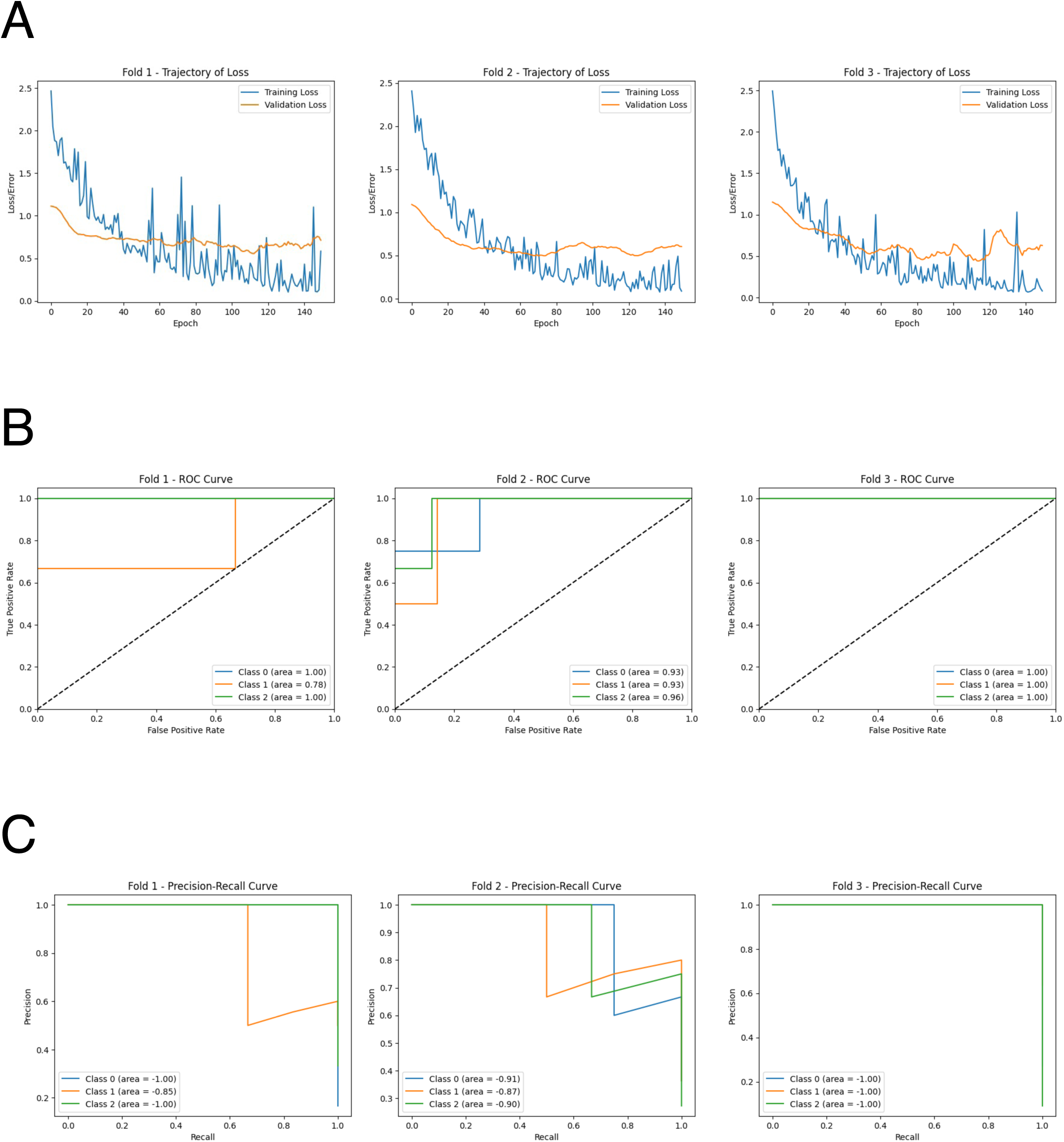
Performance of deep neural network for 3-class classification by AI. **(A)** Learning curves of the deep neural network model in 3-fold cross validation. **(B)** ROC curves of the deep neural network model in 3-fold cross validation. Class 1: CNI, class 2: AD, class 3: DLB. **(C)** Precision-recall curves of the deep neural network model in 3-fold cross validation. Class 1: CNI, class 2: AD, class 3: DLB.

### Gene Ontology analysis

We aimed to identify autoantibodies with potential pathogenic roles in dementia by conducting gene ontology analysis on the gene lists encoding the 229 autoantigens targeted by differentially elevated autoantibodies in AD and/or DLB (**Figure 4**). The analysis highlighted the “neuroactive ligand-receptor interaction” pathway in autoantibodies elevated specifically in AD. We also focused on “regulation of lipid metabolic process” highlighted only in DLB, considering recent advances in understanding the role of lipid metabolism in the pathogenesis of DLB.^51–53^

**Figure 4.**
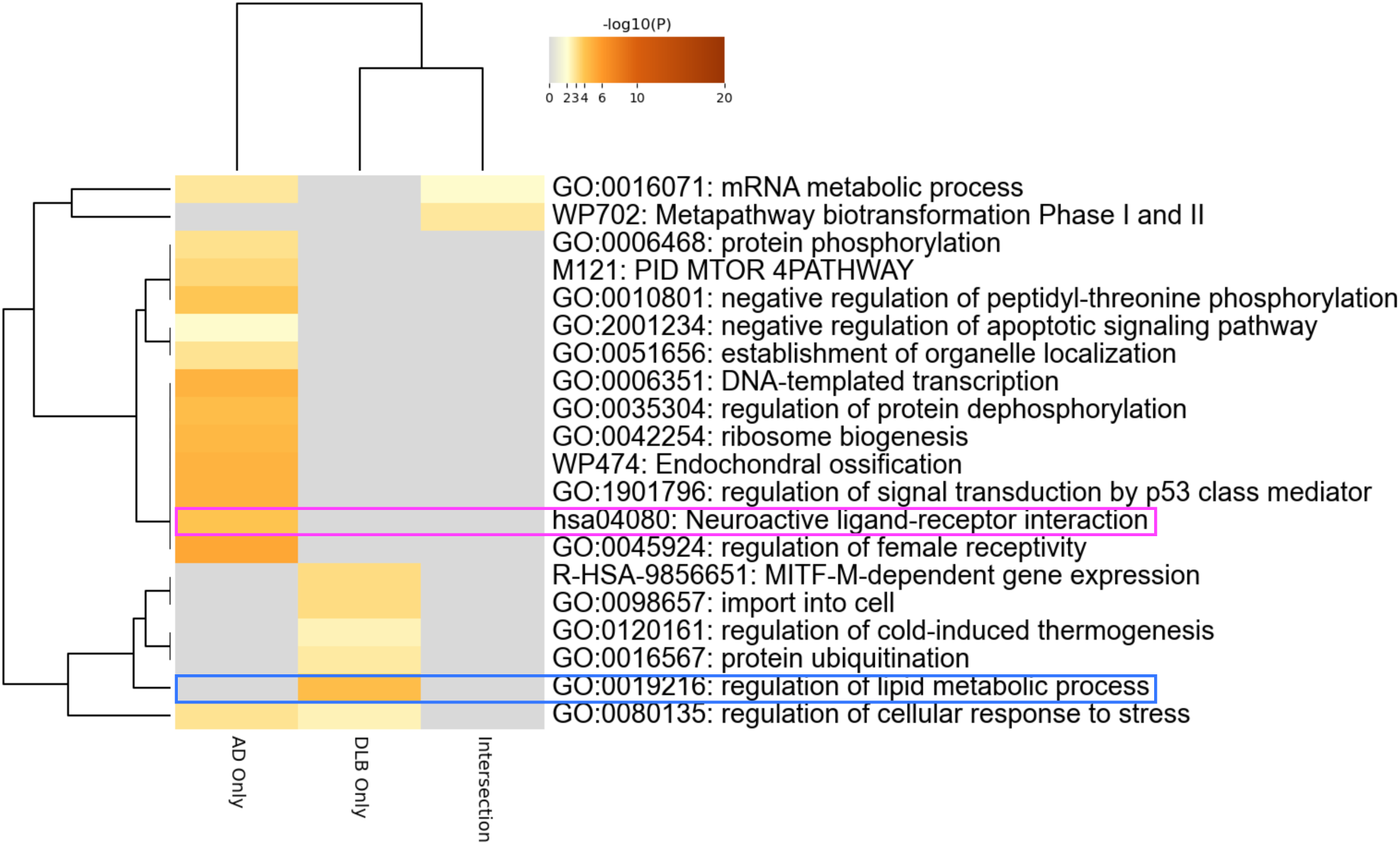
Autoantibodies to neuroactive ligand-receptor interaction-associated proteins. Gene ontology analysis encompassing the genes coding proteins targeted by autoantibodies differentially elevated in AD and/or DLB.

### Autoantibodies to neuroactive ligand-receptor interaction-associated proteins

There were 12 autoantibodies associated with neuroactive ligand-receptor interaction, and their serum levels are illustrated in **Figure 5A**. We examined the relationship between these 12 autoantibodies and clinical traits (**Figure 5B**), revealing a significant association of the serum levels of autoantibodies targeting neuropeptide B, a protein encoded by *NPB*, with female sex, presence of back pain, and MMSE scores. However, the trend of elevated serum levels of anti-NPB antibody in dementia was observed in both sex (**Extended Figure 3A**). There was no obvious cross-reactivity among the autoantibodies (**Extended Figure 3B and 3C**) and showed no disease specificity (**Extended Figure 3D**). To further investigate the potential of anti-NPB antibody to play a role in the pathogenesis of AD, we examined the correlation between serum levels of the autoantibody and all the subscales of MMSE (**Extended Figure 4**). As a result, there was statistically significant correlation in memory-related items (“Registration” and “Recall”). In line with this, a database search indicated that NPB is expressed in the CNS (**Extended Figure 5A**), including the hippocampus (**Extended Figure 5B**). The highest expression was reported in oligodendrocytes (**Extended Figure 5C**).

**Figure 5.**
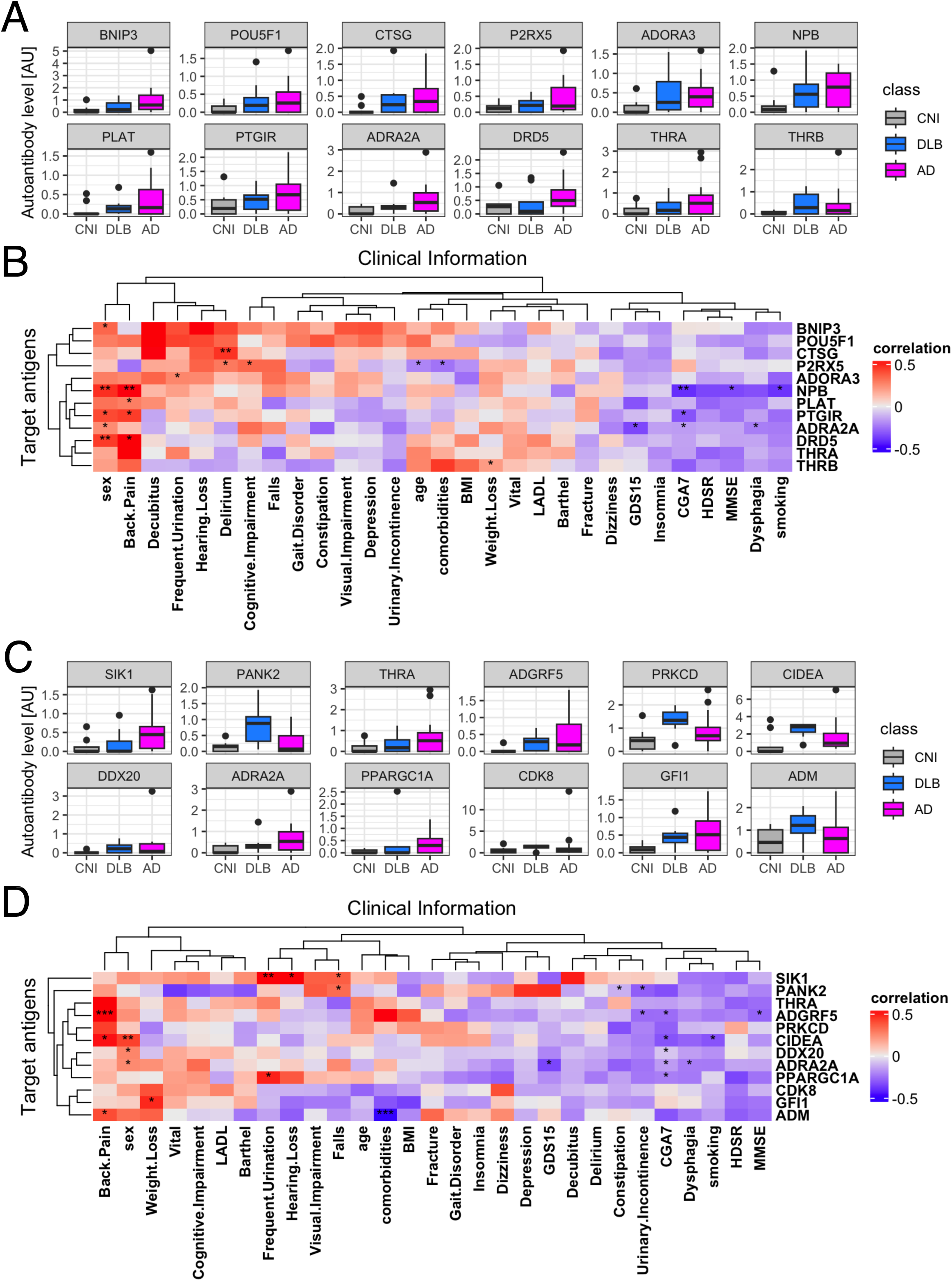
Correlation between autoantibodies highlighted in gene ontology analysis and clinical traits of dementia. **(A)** Box plots describe the serum levels of autoantibodies to neuroactive ligand-receptor interaction-associated proteins. **(B)** Heatmap illustrates correlation between autoantibodies to neuroactive ligand-receptor interaction-associated proteins and demographic and clinical characteristics of dementia. **(C)** Box plots describe the serum levels of autoantibodies to regulation of lipid metabolic process-associated proteins. **(D)** Heatmap illustrates correlation between autoantibodies to regulation of lipid metabolic process-associated proteins and demographic and clinical characteristics of dementia. *: P < 0.05, **: P < 0.01. P values were calculated by Spearman’s correlation test.

### Autoantibodies to lipid metabolism-associated proteins

Finally, we focused on 12 autoantibodies targeting lipid metabolism-associated proteins, whose serum levels are illustrated in **Figure 5C**. We examined the relationship between these 12 autoantibodies and clinical traits (**Figure 5D**). This analysis revealed a significant association of the serum levels of autoantibodies targeting Adhesion G Protein-Coupled Receptor F5 (ADGRF5) encoded by *ADGRF5* with presence of back pain, lower Comprehensive Geriatric Assessment 7 (CGA7) scores, and lower MMSE scores, especially in “Registration” and “Repetition” subscales (**Extended Figure 6**). There was no big difference between both sex (**Extended Figure 7A**), cross-reactivity, nor disease specificity. (**Extended Figure 7B, 7C, and 7D**). A database search indicated that the expression of ADGRF5 is ubiquitous across various human tissues (**Extended Figure 8A**), including the CNS (**Extended Figure 8B**), predominantly in microglial cells (**Extended Figure 8C**).

## Discussion

In this study, we utilized our proprietary PWAS technique to analyze serum samples from patients with AD, DLB, and CNIs. Our results showed an increase in the overall levels of autoantibodies in AD and DLB patients compared to CNIs (**Figure 1A**). We identified 229 autoantibodies that were differentially elevated in AD and/or DLB (**Figure 1D**), effectively distinguishing between AD, DLB, and CNI groups (**Figure 1F**). Machine learning applied to these 229 autoantibodies demonstrated high accuracy in differentiating AD patients from others (**Table 1**), and even achieved success in multi-class classification (**Figure 3**). Gene ontology analysis highlighted autoantibodies targeting neuroactive ligands and receptors in AD, including anti-NPB antibody, as well as lipid metabolism-associated proteins in DLB, such as anti-ADGRF5 antibody (**Figure 4**). Both of anti-NPB and anti-ADGRF5 autoantibodies showed significant correlation with total MMSE scores (**Figure 5B and 5D**) and memory-related subscale scores (**Extended Figure 4 and 6**). Considering the expression of NPB and ADGRF5 in the central nervous system (**Extended Figure 5 and 8**), these findings suggest that autoantibodies targeting NPB or ADGRF5 may contribute to the pathogenesis of dementia. Our results underscore the potential of our systems-based approach in developing novel diagnostic tools and propose a new research strategy to explore the autoimmune aspects of dementia.

A key highlight of our analysis is the ability of AI integrated with our multiplex autoantibody measurement to achieve near-perfect accuracy in classifying AD versus other groups (**Table 1**) and even in multi-class classification tasks (AD, DLB, and CNI; **Figure 3**). This concept has already been demonstrated in other autoimmune and malignant disorders,^47,48^ and is partially available commercially as the Autoantibody Array Assay (A-Cube).^44^ Given that blood tests are less invasive than other procedures like cerebrospinal fluid collection and radiological imaging studies and can be conducted without causing undue concern to the patient about suspected cognitive impairment, multiplex measurement of serum autoantibodies using WPAs and AI-based interpretation represents a promising strategy for diagnosing dementia and its subtypes.

The *NPB* gene encodes neuropeptide B, a short biologically active peptide that acts as an agonist for GPCRs known as neuropeptide B/W receptors 1 (NPBWR1) and 2 (NPBWR2).^54^ Neuropeptide B is believed to play roles in regulating feeding, the neuroendocrine system, memory, learning, and the pain pathway.^55^ Research by Nagata-Kuroiwa R *et al.* on NPBWR1 knockout mice revealed increased autonomic and neuroendocrine responses to physical stress and abnormalities in contextual fear conditioning, suggesting a role for NPBWR1 in stress vulnerability and fear memory.^56^ Histological and electrophysiological studies indicate that NPBWR1 acts as an inhibitory regulator on a subpopulation of GABAergic neurons in the lateral division of the central nucleus of the amygdala, terminating stress responses. Additionally, Watanabe N *et al.* demonstrated that a single nucleotide polymorphism in NPBWR1, associated with impaired molecular function, affected valence evaluation and dominance ratings in response to seeing angry faces in humans, suggesting NPBWR1’s involvement in social interaction.^57^ These insights highlight the potential role of autoantibodies affecting the NPB-NPBWR1 signaling system in social behavior, suggesting its potential contribution to the clinical manifestations of AD, particularly its behavioral and psychological symptoms.

Our study also revealed a strong association between serum anti-NPB antibody levels and the presence of back pain, likely due to the role of NPB-NPBWR1 signaling in pain transmission. NPB knockout mice exhibit different responses to pain; they show hyperalgesia to acute inflammatory pain but not to thermal or chemical pain.^58^ Intrathecal administration of NPB reduced mechanical allodynia via activation of NPBWR1 receptors without affecting thermal hyperalgesia.^59^ These effects were not inhibited by naloxone, an opioid receptor antagonist, indicating the involvement of a non-opioid analgesic pathway, possibly related to myelin-forming Schwann cells, which express low levels of NPBWR1 under physiological conditions but much higher levels in patients with inflammatory neuropathies. Thus, anti-NPB antibodies may play a role in modulating nociceptive transmission.

ADGRF5, a member of the adhesion GPCR (aGPCR) family, which is the second largest GPCR subfamily, has recently garnered attention for its biological functions, disease relevance, and potential as a drug target.^60^ Predominantly expressed in the lung and kidney, ADGRF5 may play a crucial role in regulating surfactant protein synthesis acid-base balance in these organs.^61–63^ DiBlasi *et al.* identified a single nucleotide polymorphism in the ADGRF5 gene linked to an increased risk of suicide,^64^ suggesting its psychiatric role. Additionally, Kaur *et al.* found that plasma levels of ADGRF5 are associated with the APOE genotype,^65^ a known risk factor for DLB and AD.^52,53^ Elevated levels of anti-ADGRF5 antibodies correlated with global geriatric function scores assessed by CGA7 (**Figure 5D**), and the fact that ADGRF5 expression is not exclusive to the CNS (**Extended Figure 8**), may reflect systemic aspects of DLB affecting multiple organs.^66^

It is important to note that not all patients had anti-NPB nor anti-ADGRF5 antibodies, and their serum levels in AD were not specific to the condition (**Extended Figure 3D and 7D**). This suggests that while the presence of these autoantibodies may not explain the entire pathogenesis of dementia, they could influence disease manifestation and progression as bystanders. Further investigation is needed to clarify the role of anti-NPB and anti-ADGRF5 antibodies in the pathophysiology, including functional assays to assess the effects of these antibodies on neurons or glial cells, passive immune challenge in AD animal models by administering anti-NPB or anti-ADGRF5 antibodies, and active immunization of animals with NPB or ADGRF5 antigens.

Our study has several strengths. First, by including multiple types of dementia (AD and DLB), as well as CNIs, we were able to identify autoantibodies that are differentially elevated in each condition and develop machine learning methodologies for distinguishing different types of dementia in a relatively non-invasive way. Second, the use of a wheat-germ *in vitro* protein synthesis system and the manipulation technique for WPAs allowed for high-throughput expression of a wide range of human proteins, including soluble proteins, on a single platform.^41,42,67^ This enabled our autoantibody measurement to cover an almost proteome-wide range of antigens, allowing the application of omics-based bioinformatics approaches to interpret the data. Third, integration of AI and omics-based approach allowed us to conduct an unbiased and holistic investigation, resulting in novel discoveries.

A major limitation of our study is the demographic differences among the human subjects, particularly in terms of sex (**Extended Table 1**). Moreover, the sample size was modest, lacked external validation, and was cross-sectional. Future studies should target larger, more demographically balanced patient groups with a wider range of dementia types, such as VD and FTD. Recruiting longitudinal specimens and data from elderly individuals before and after the onset of MCI in prospective population-based cohorts would be a valuable challenge to explore the causal relationship between autoantibodies and dementia pathogenesis.

## Data Availability

All data produced in the present study are available upon reasonable request to the authors.

## Acknowledgements

We thank Ms. Maiko Enomoto and her colleagues for their secretarial work. We appreciate K. Yamaguchi, T. Okumura, C. Ono, A. Sato, A. Miya, and N. Goshima from ProteoBridge Corporation for preparing the WPAs. We also acknowledge R. Uchino, Y. Murakami, and H. Matsunaka from TOKIWA Pharmaceuticals Co. Ltd. for providing technical assistance with autoantibody measurement.

## Author Contributions

KM Matsuda primarily engaged in autoantibody measurement, data analysis, visualization, and writing the first draft of the manuscript. Y Umeda-Kameyama oversaw clinical sample and data collection and was involved in revising the manuscript. K Iwadoh participated in machine learning analysis. M Miyawaki, M Yakabe, S Ogawa, and M Akishita participated in clinical sample and information collection. S Sato conceptualized and supervised the study. A Yoshizaki conceptualized, launched, and supervised this study, and was involved in revising the manuscript.

## Conflict-of-interest statement

A Yoshizaki belongs to the Social Cooperation Program, Department of Clinical Cannabinoid Research, The University of Tokyo Graduate School of Medicine, Tokyo, Japan, supported by Japan Cosmetic Association and Japan Federation of Medium and Small Enterprise Organizations. The remaining authors declare that the research was conducted in the absence of any commercial or financial relationships that could be construed as a potential conflict of interest.

## Materials and Methods

### Participants

We enrolled 26 dementia participants who were admitted to the Department of Geriatric Medicine, The University of Tokyo Hospital, Tokyo, Japan, for evaluation of cognitive impairment. All participants were diagnosed by experienced geriatricians using DSM-IV criteria for AD (n=18), and Revised 2017 Clinical Diagnostic Criteria for Dementia with Lewy Bodies for DLB (n=8).^35^ Nine participants were NCIs who admitted to the Department of Geriatric Medicine, The University of Tokyo Hospital, for other reasons, except acute illness and autoimmune disease. Patients with malignant disorders were excluded. We made precise diagnoses using psychological tests, information from family, laboratory data, brain structural imaging (X-ray computed tomography or nuclear magnetic resonance imaging). We also used N-isopropyl-p-iodoamphetamine brain perfusion single photon emission computed tomography (SPECT), metaiodobenzylguanidine, ioflupane dopamine transporter SPECT. Clinical metrics included number of comorbidities, Charlson’s Comorbidity Index, Comprehensive Geriatric Assessment-short version (CGA7), MMSE, HSDR, Barthel Index, Lowton’s Instrumental Activities of Daily Living (IADL) scores, Geriatric Depression Scale 15 (GDS15), and Vitality Index. All procedures were approved by the Ethical Review Board at The University of Tokyo Hospital and The University of Tokyo (approval number 2797). The clinical study guidelines of the University of Tokyo, which conform to the Declaration of Helsinki, were strictly adhered to CNIs, dementia patients and their families. They were provided with detailed information about the study, and all provided written informed consent to participate.

### Autoantibody measurement

WPAs were arranged as previously described.^43^ First, proteins were synthesized *in vitro* utilizing a wheat germ cell-free system from 13,455 clones of the HuPEX.^41^ Second, synthesized proteins were plotted onto glass plates (Matsunami Glass, Osaka, Japan) in an array format by the affinity between the GST-tag added to the N-terminus of each protein and glutathione modified on the plates. The WPAs were treated with human serum diluted by 3:1000 in the reaction buffer containing 1x Synthetic block (Invitrogen), phosphate-buffered saline (PBS), and 0.1% Tween 20. Next, the WPAs were washed, and goat anti-Human IgG (H+L) Alexa Flour 647 conjugate (Thermo Fisher Scientific, San Jose, CA, USA) diluted 1000-fold was added to the WPAs and reacted for 1 hour at room temperature. Finally, the WPAs were washed, air-dried, and fluorescent images were acquired using a fluorescence imager (Typhoon FLA 9500, Cytiva, Marlborough, MA, USA). Fluorescence images were analyzed to quantify serum levels of autoantibodies targeting each antigen, following the formula shown below:

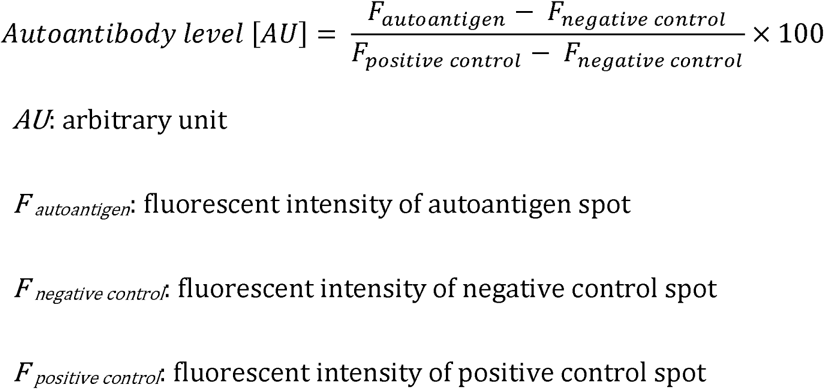

### AI-based analysis

We applied supervised machine learning techniques using Python (v3.10.12) with libraries from Scikit-learn and the PyTorch framework to construct classifiers for the diagnosis of dementia based on the autoantibody measurement data. The performance of the classifiers was evaluated with 3-fold cross validation, using the metrics of area under the receiver operating characteristics curve (AUC), area under the precision-recall curve, accuracy, precision, recall, and F1-score, with the higher score indicating the better classification performance. Machine learning models from Scikit-learn included simple linear regression, Lasso regression, Ridge regression, logistic regression, support vector machine (SVM), random forest, XGBoost, LightGBM, CatBoost, decision trees, gradient boosting machines and naïve Bayes to conduct binary classification. Additionally, we also used deep neural networks with five hidden layers in PyTorch to classify between three types of dementia.

### Statistical analysis

Fisher’s exact test was performed to compare categorical variables. Mann-Whitney U test was performed to compare continuous variables. Spearman correlation test was used for correlation analysis. P values of < 0.05 were considered statistically significant. Data analyses were conducted using R (v4.2.1).

### Protein functional enrichment analysis

Gene Ontology Analysis using web-based tools targeted the list of the entry clones coding the differentially highlighted autoantigens was performed for gene-list enrichment analysis, gene-disease association analysis, and transcriptional regulatory network analysis with Metascape.^68^

### Sequence identity analysis

To assess cross-reactivity among proteins that express similar antigen epitopes and are highly correlated, we checked the correlation of the differentially expressed autoantibodies. The corresponding proteins of the highly correlated autoantibodies (Spearman’s r > 0.5) were then aligned with the highly correlated proteins using the Uniprot alignment tool.

### Data visualization

Box plots, scatter plots, hierarchical clustering, and correlation matrix were visualized by using R (v4.2.1). Box plots were defined as follows: the middle line corresponds to the median; the lower and upper hinges correspond to the first and third quartiles; the upper whisker extends from the hinge to the largest value no further than 1.5 times the interquartile range (IQR) from the hinge; and the lower whisker extends from the hinge to the smallest value at most 1.5 times the IQR of the hinge.

**Extended Figure 1. Sum of autoantibody levels by age and sex. (A)** Box plots show SAL by age groups. **(B)** Box plots show SAL by sex.

**Extended Figure 2. Additional information for autoantibodies highlighted in 2-class classification tasks by AI. (A)** Box plots describe the serum levels of autoantibodies highlighted in 2-class classification tasks by sex. **(B)** A correlation matrix of the autoantibodies highlighted in 2-class classification tasks using Spearman’s correlation. Only statistically significant pairs (P < 0.05) are shown. **(C)** Identity matrix, generated from aligning the corresponding protein sequences of the highly correlated autoantibodies (Spearman’s r > 0.5). **(D)** Box plots describe the serum levels of autoantibodies highlighted in 2-class classification tasks in COVID-19, atopic dermatitis, anti-neutrophil cytoplasmic antibody-associated vasculitis, systemic lupus erythematosus, systemic sclerosis, and healthy controls. The data derives from the UT-ABCD.

**Extended Figure 3. Additional information for autoantibodies to neuroactive ligand-receptor interaction-associated proteins. (A)** Box plots describe the serum levels of autoantibodies to neuroactive ligand-receptor interaction-associated proteins by sex. **(B)** A correlation matrix of the autoantibodies to neuroactive ligand-receptor interaction-associated proteins using Spearman’s correlation. Only statistically significant pairs (P < 0.05) are shown. **(C)** Identity matrix, generated from aligning the corresponding protein sequences of the highly correlated autoantibodies (Spearman’s r > 0.5). **(D)** Box plots describe the serum levels of autoantibodies to neuroactive ligand-receptor interaction-associated proteins in COVID-19, atopic dermatitis, anti-neutrophil cytoplasmic antibody-associated vasculitis, systemic lupus erythematosus, systemic sclerosis, and healthy controls. The data derives from the UT-ABCD.

**Extended Figure 4. Correlation between serum levels oof anti-NPB antibodies and MMSE subscales.**

**Extended Figure 5. Expression of the *NPB* gene in human tissues and single cells. (A)** Expression of *NPB* in multiple human tissues measured by bulk RNA-sequencing from the Human Protein Atlas. **(B)** Expression of *NPB* in the *CNS* from the Human Protein Atlas. **(C)** Expression of *NPB* in the CNS evaluated by single-cell RNA-sequencing from the Human Protein Atlas.

**Extended Figure 6. Correlation between serum levels oof anti-ADGRF5 antibodies and MMSE subscales.**

**Extended Figure 7. Additional information for autoantibodies to regulation of lipid metabolic process-associated proteins. (A)** Box plots describe the serum levels of autoantibodies to regulation of lipid metabolic process-associated proteins by sex. **(B)** A correlation matrix of the autoantibodies to regulation of lipid metabolic process-associated proteins using Spearman’s correlation. Only statistically significant pairs (P < 0.05) are shown. **(C)** Identity matrix, generated from aligning the corresponding protein sequences of the highly correlated autoantibodies (Spearman’s r > 0.5). **(D)** Box plots describe the serum levels of autoantibodies to regulation of lipid metabolic process-associated proteins in COVID-19, atopic dermatitis, anti-neutrophil cytoplasmic antibody-associated vasculitis, systemic lupus erythematosus, systemic sclerosis, and healthy controls. The data derives from the UT-ABCD.

**Extended Figure 8. Expression of the *ADGRF5* gene in human tissues and single cells. (A)** Expression of *ADGRF5* in multiple human tissues measured by bulk RNA-sequencing from the Human Protein Atlas. **(B)** Expression of *ADGRF5* in the *CNS* from the Human Protein Atlas. **(C)** Expression of *ADGRF5* in the CNS evaluated by single-cell RNA-sequencing from the Human Protein Atlas.

## Notes

### Funding Statement

This study did not receive any funding.

### Author Declarations

All procedures were approved by the Ethical Review Board at The University of Tokyo Hospital and The University of Tokyo (approval number 2797).

