## Supplementary figures and images for "Artificial intelligence and omics-based autoantibody profiling highlights autoimmunity targeting ligand-receptor interaction in dementia"

### Extended Figure 1

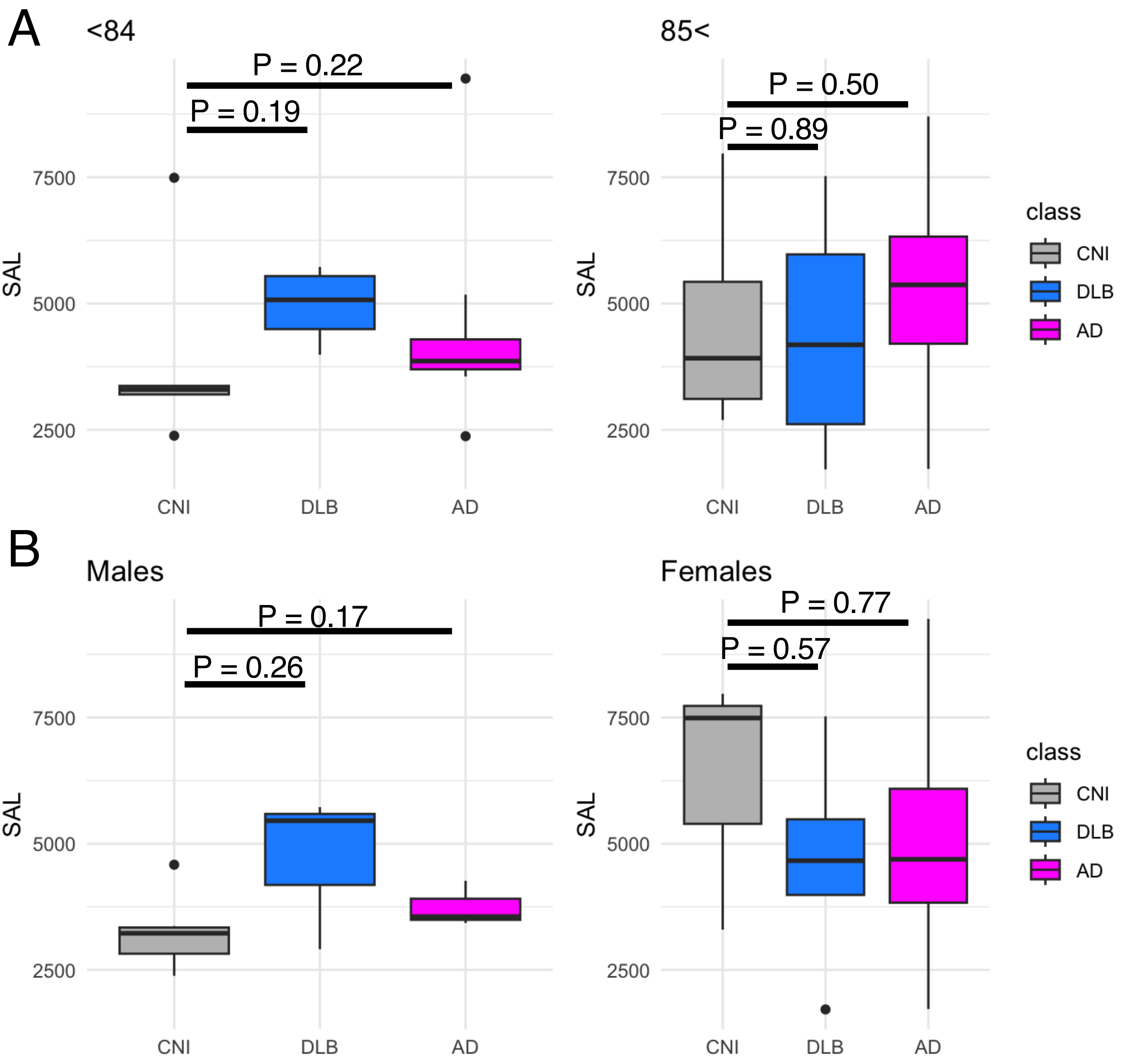


Matsuda KM et al.
Extended Figure 1

### Extended Figure 2

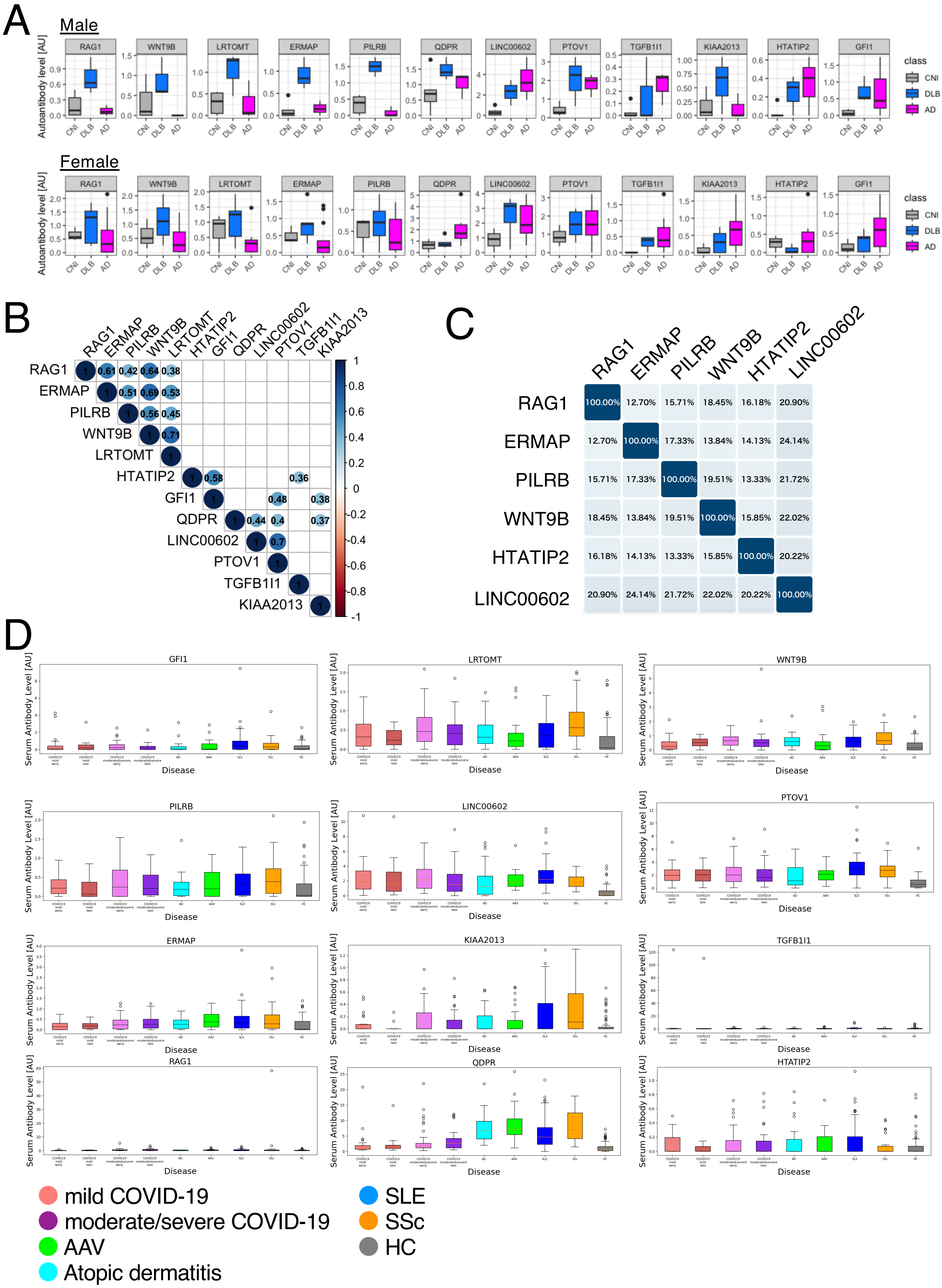


Matsuda KM et al.
Extended Figure 2

### Extended Figure 3

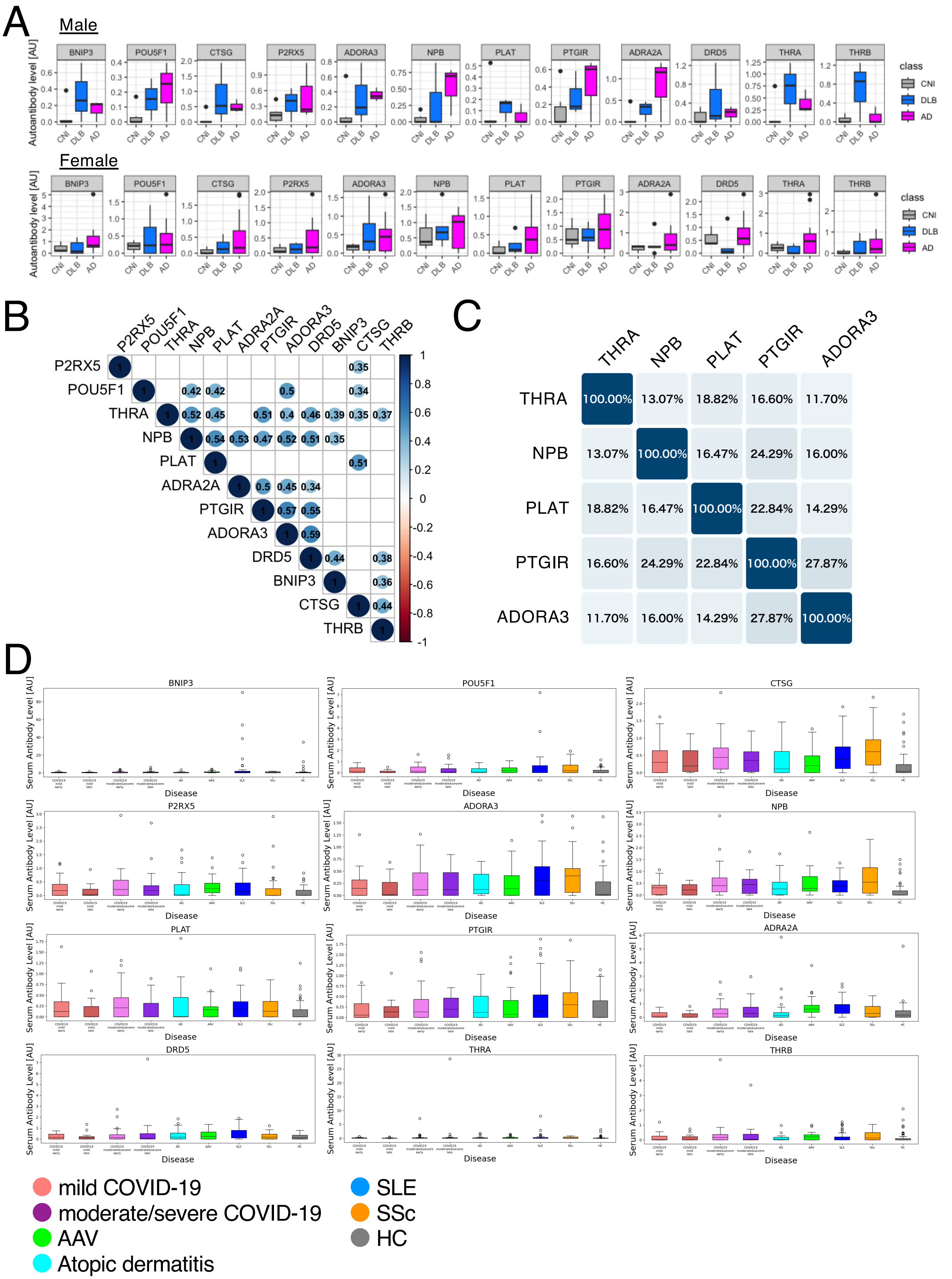


Matsuda KM et al.
Extended Figure 3

### Extended Figure 4

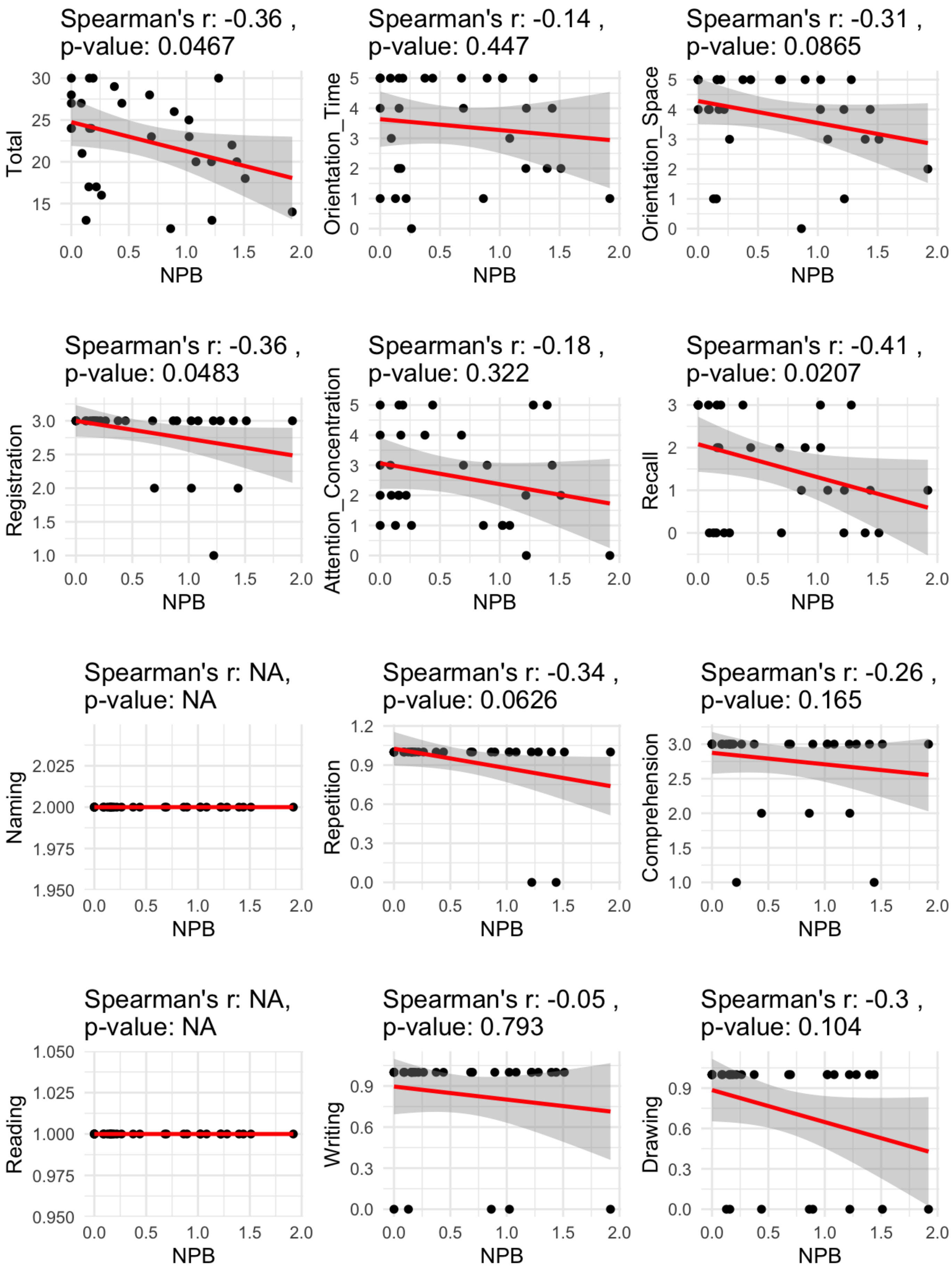


Matsuda KM et al.
Extended Figure 4

### Extended Figure 5

A

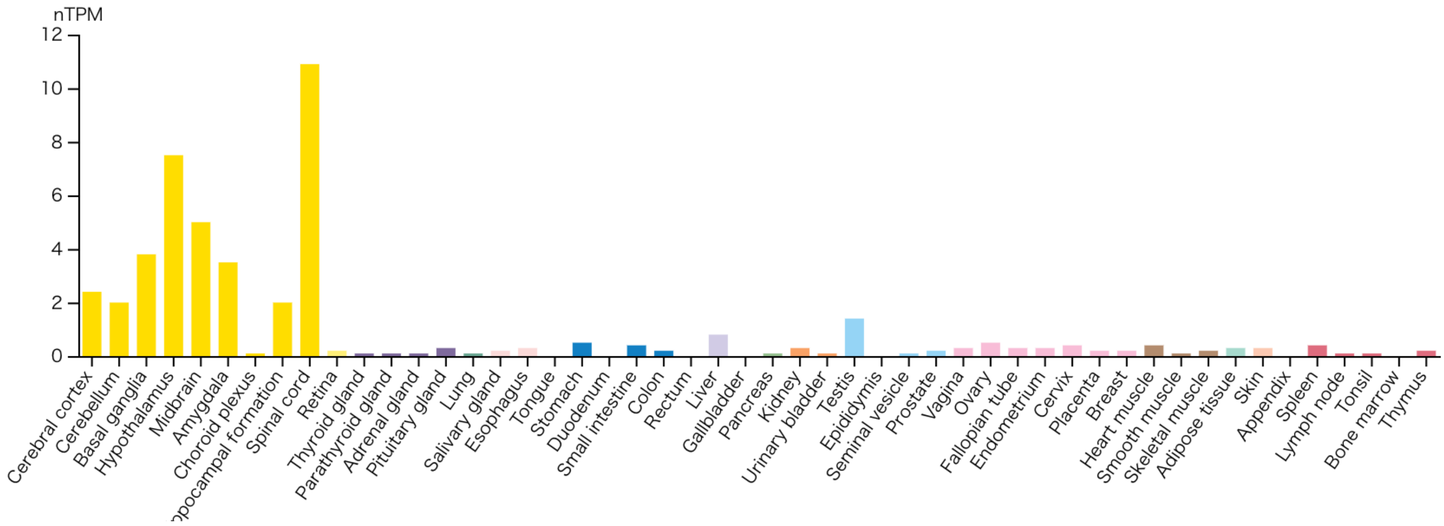


B

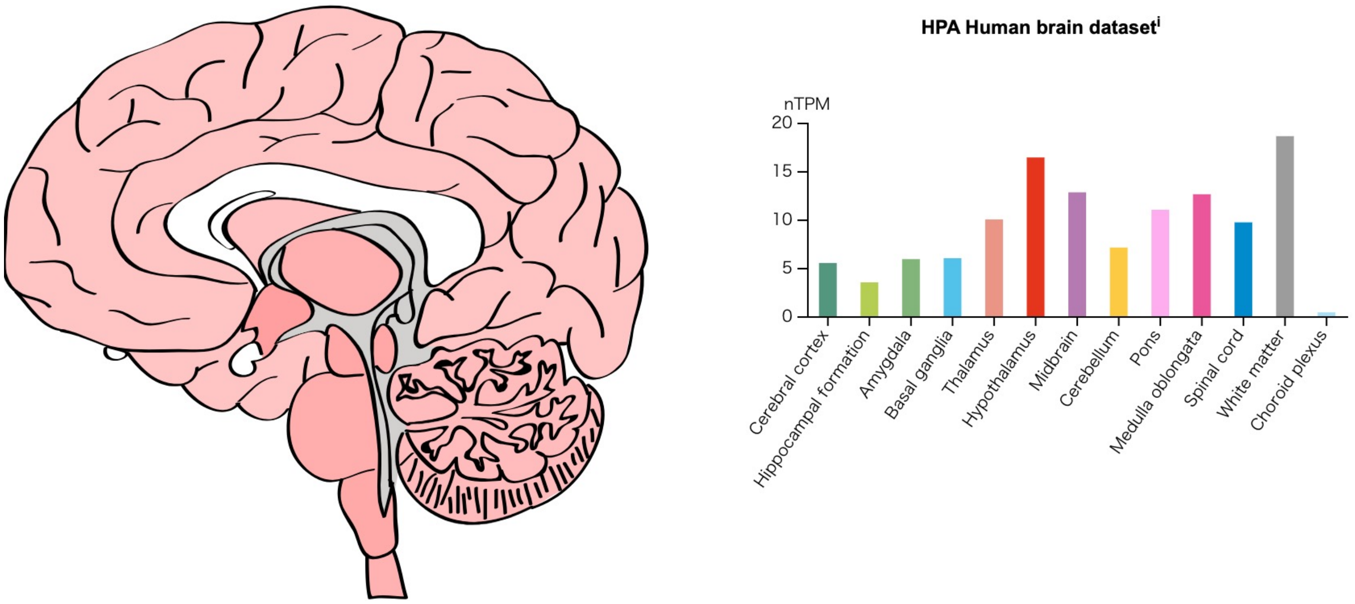


C

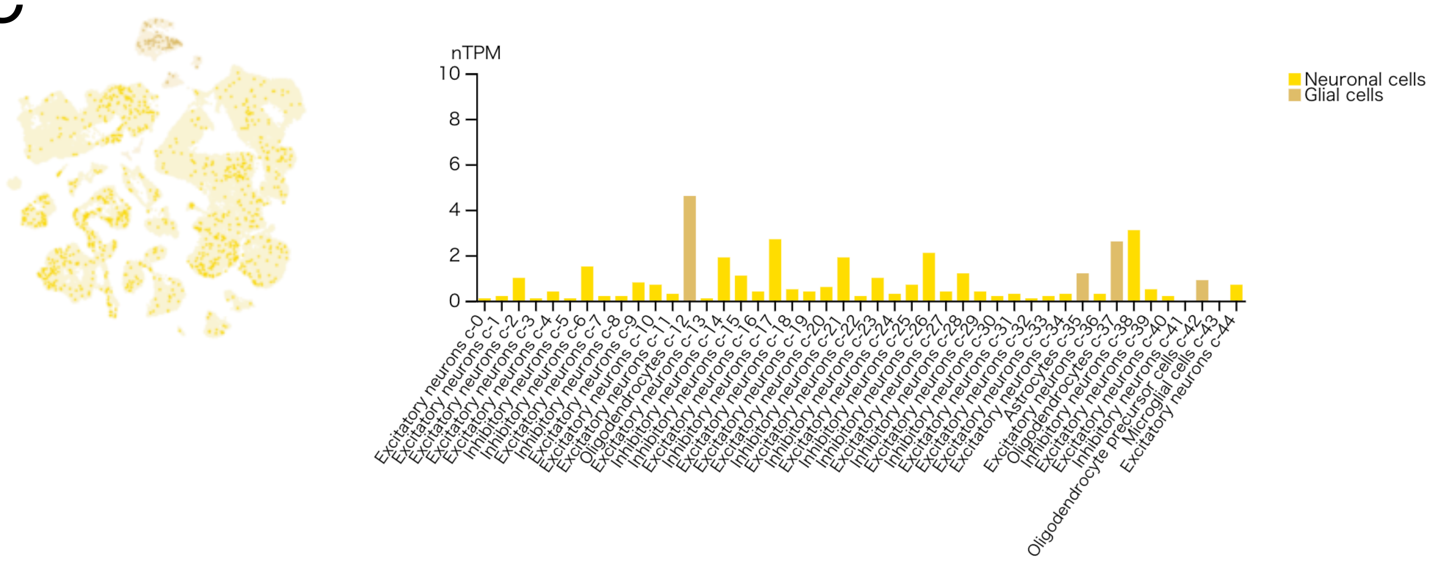


Matsuda KM et al.
Extended Figure 5

### Extended Figure 6

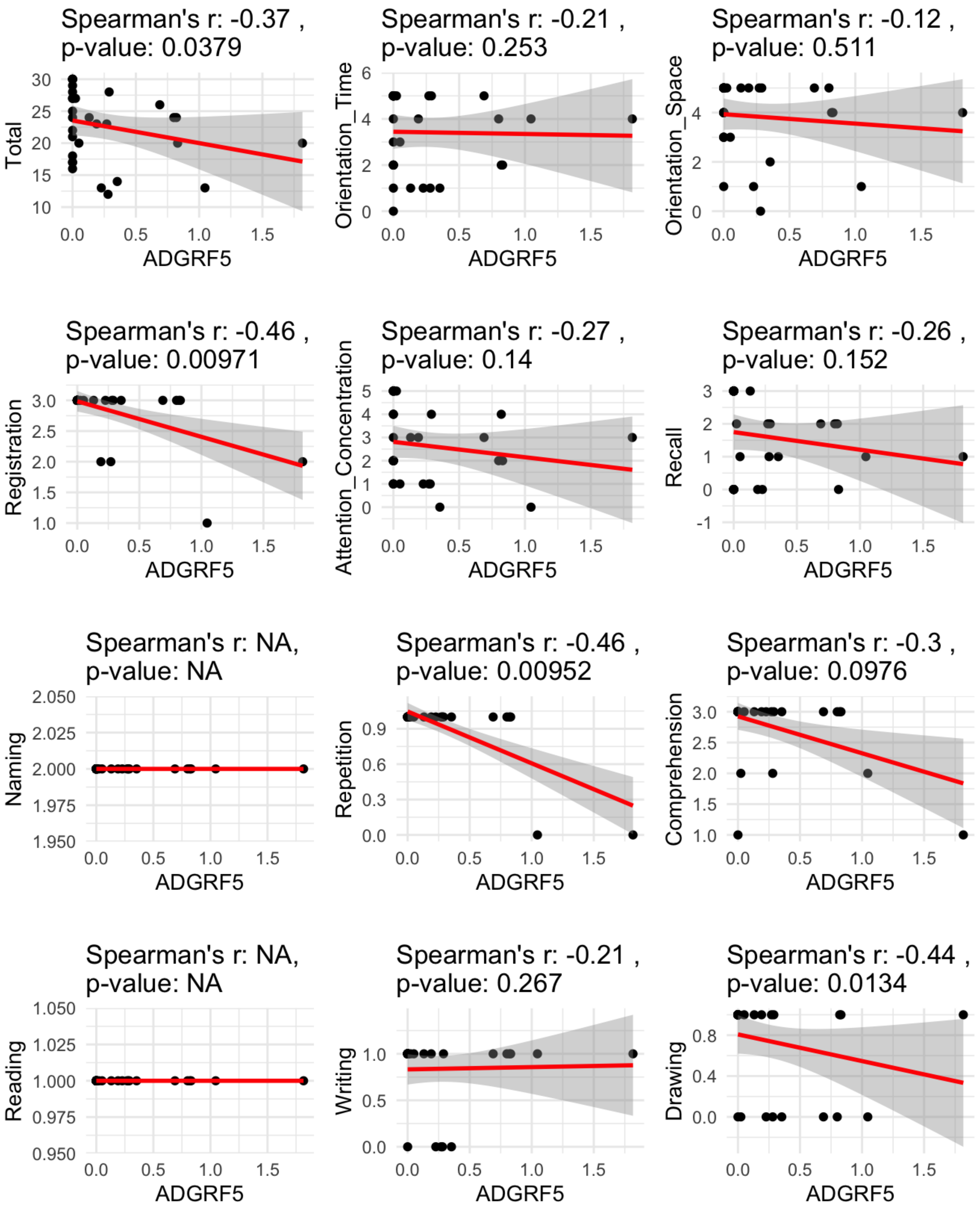


Matsuda KM et al.
Extended Figure 6

### Extended Figure 7

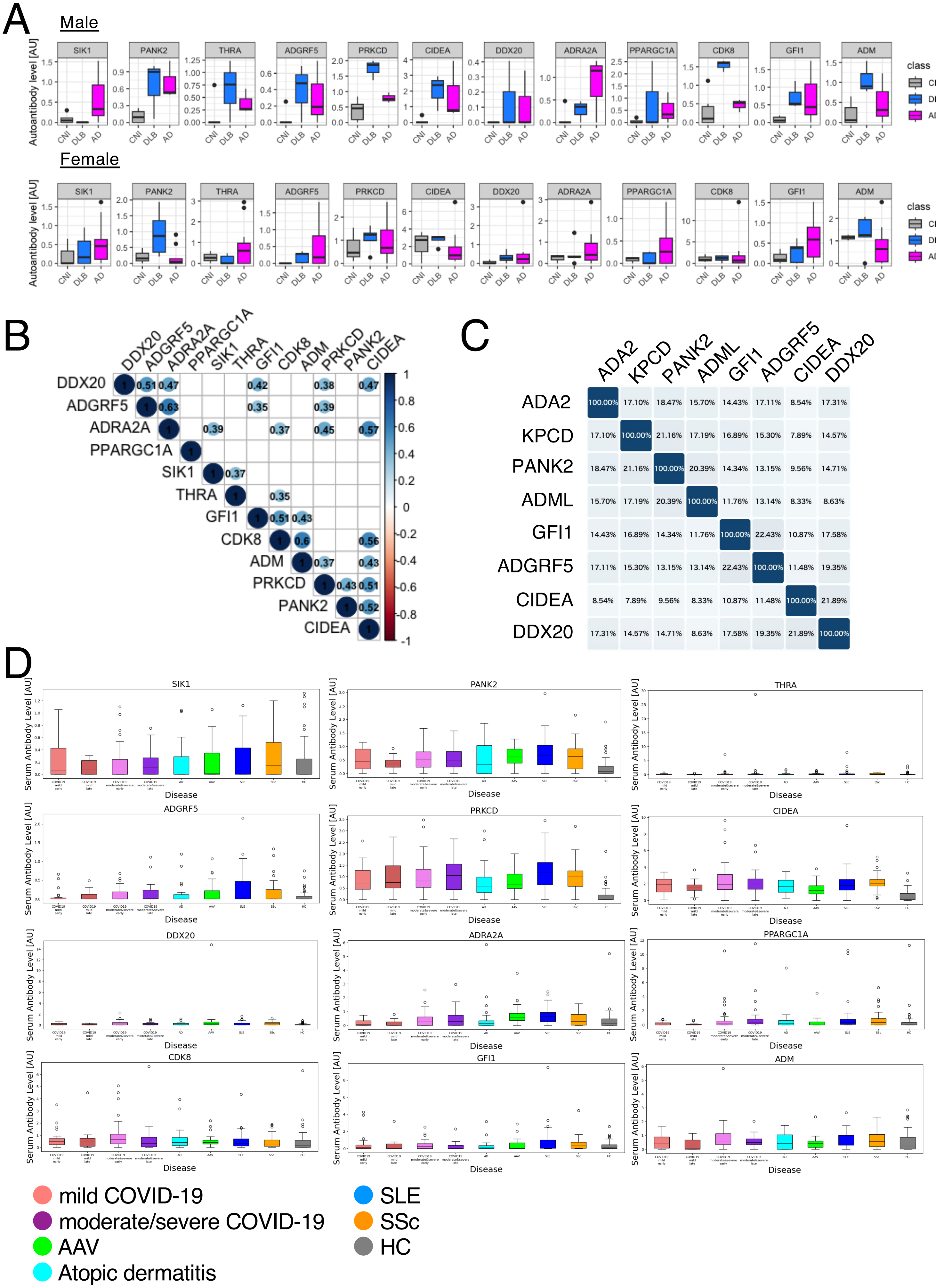


Matsuda KM et al.
Extended Figure 7

### Extended Figure 8

A

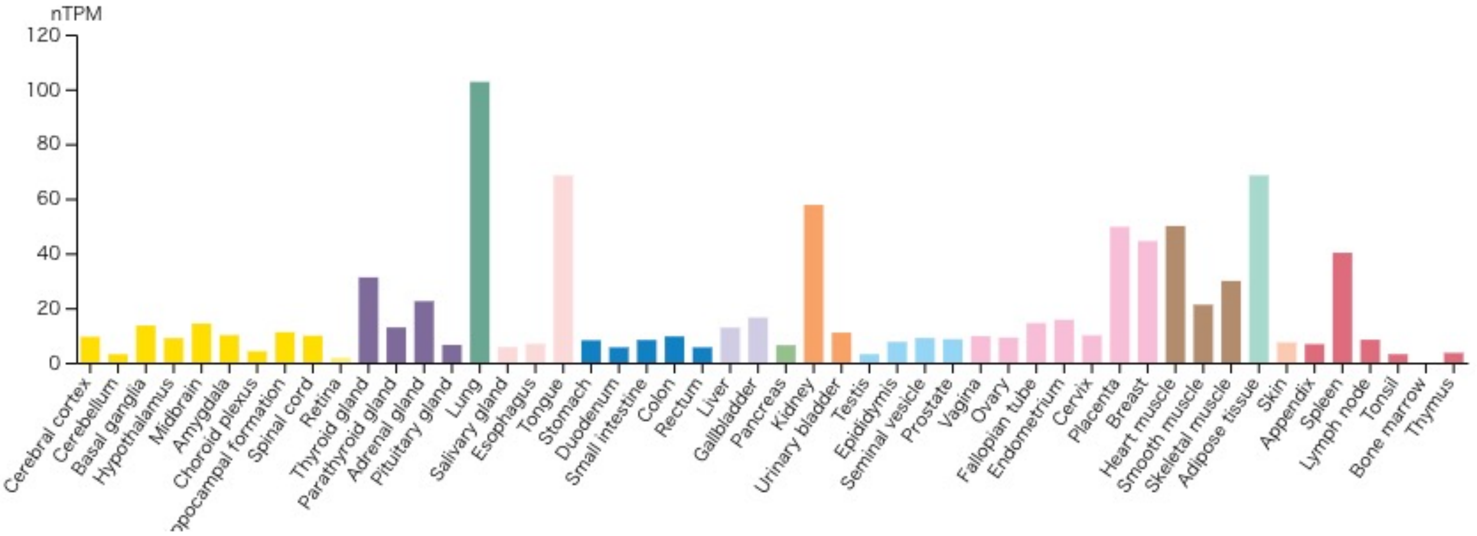


B

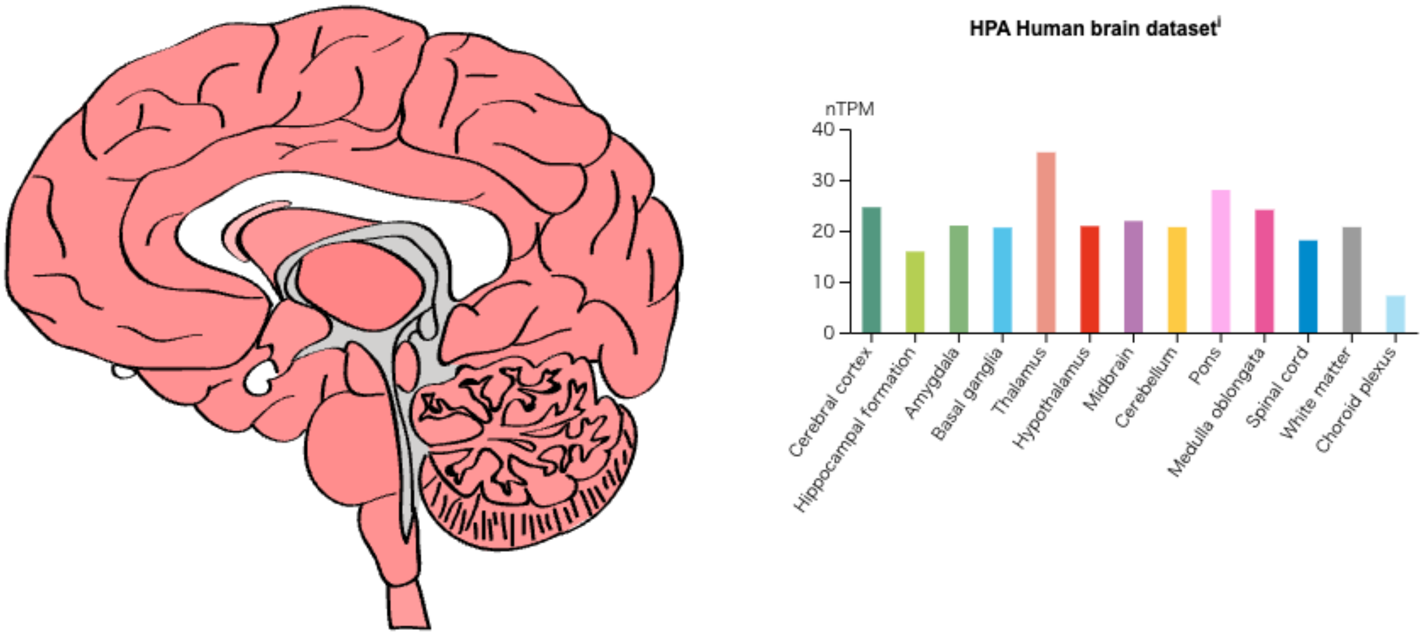


C

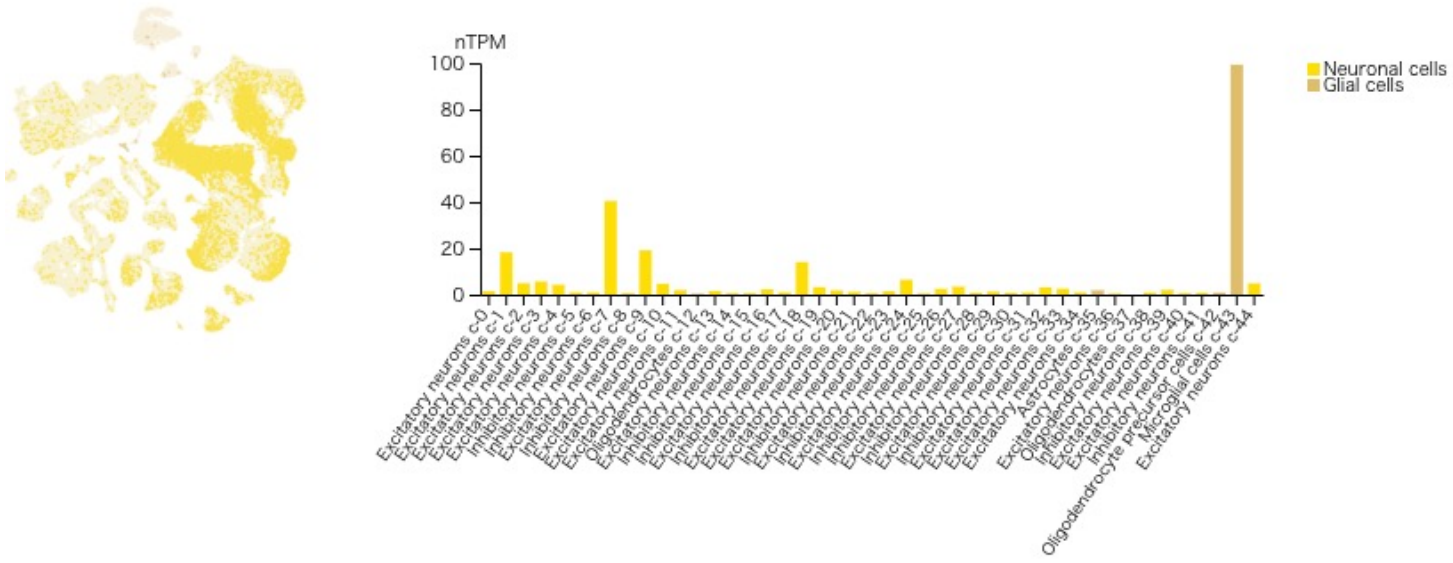


Matsuda KM et al.
Extended Figure 8
